# Quantifying Ventricular CSF Clearance in the Human Brain Using Dynamic 18F-FDG PET: Insights into Age-Related Glymphatic Impairment

**DOI:** 10.1101/2025.04.07.25325241

**Authors:** Zeyu Zhou, Tianyun Zhao, John Gardus, Qiuting Wen, Yang Feng, Christine DeLorenzo, Ramin Parsey, Chuan Huang

**Affiliations:** Department of Radiology and Imaging Sciences, Emory University School of Medicine; Department of Biomedical Engineering, Stony Brook University; Department of Psychiatry and Behavior Sciences, Stony Brook University; Department of Radiology and Imaging Sciences, Indiana University School of Medicine; Department of Biostatistics, New York University School of Global Public Health; Department of Biomedical Engineering, Georgia Institute of Technology and Emory University

## Abstract

**Purpose:** The glymphatic system facilitates brain waste clearance via cerebrospinal fluid (CSF) flow, and its dysfunction has been linked to aging and neurodegeneration. However, clinically accessible methods to quantify glymphatic function in humans remain limited. This study aimed to examine the potential of dynamic 18F-FDG PET for measuring ventricular CSF clearance - as a surrogate marker of glymphatic function. Specifically, we evaluated its association with age, its test–retest reliability, and the feasibility of reduced scan durations for clinical applicability.

**Methods:** We analyzed 72 baseline 18F-FDG PET scans from participants enrolled in a prior depression trial. Time–activity curves (TACs) were extracted from the lateral ventricles and fitted with a γ-variate model to estimate influx (*μ*_*in*_) and clearance (*μ*_*out*_) parameters. Associations with age and clinical factors were examined using correlation and multiple linear regression. Test–retest reliability was assessed in 11 placebo-treated participants who underwent repeat scans eight weeks apart. A feasibility analysis tested whether shorter scan windows could yield comparable clearance estimates.

**Results:** *μ*_*out*_ showed a strong negative correlation with age (r = –0.680, p < 0.001), while *μ*_*in*_ was not significantly age-related. Age remained a significant predictor of *μ*_*out*_ after adjusting for sex, ventricle size, and depression severity. A positive association between *μ*_*out*_ and depression severity was observed after covariate adjustment. Test–retest analysis yielded an intraclass correlation coefficient of 0.702 for *μ*_*out*_, indicating moderate-to-good reproducibility. A shortened 30-minute scan window (starting 30 minutes post injection) preserved strong correlations with both *μ*_*out*_ and age, supporting the potential for abbreviated imaging protocols.

**Conclusion:** Dynamic 18F-FDG PET provides a reliable and noninvasive method to quantify ventricular CSF clearance, revealing age-related decline indicative of glymphatic impairment. The method demonstrates reproducibility over time and retains key clearance metrics even with shortened scan durations. These findings establish a clinically feasible 18F-FDG PET-based approach for studying brain clearance and glymphatic function in aging and disease.

## Introduction

The glymphatic system is a waste clearance pathway in the brain that plays a crucial role in maintaining neural homeostasis by facilitating the exchange of cerebrospinal fluid (CSF) and interstitial fluid (ISF) through perivascular spaces. Dysfunction of this system has been implicated in a range of neurodegenerative disorders, including Alzheimer’s disease (AD), Parkinson’s disease (PD), and vascular dementia, where impaired clearance of metabolic waste may contribute to disease progression ^1,2^. In addition, glymphatic dysfunction has been linked to cerebrovascular and traumatic brain disorders, such as small vessel disease^3^, traumatic brain injury^4^, stroke-related impairment^5^, and idiopathic normal pressure hydrocephalus^6^. CSF flow is a fundamental component of glymphatic function, serving as the driving force behind solute transport and waste removal from the brain^7,8^. In parallel with pharmacological and lifestyle interventions, novel surgical procedures such as lymphovenous anastomosis (LVA) have been proposed to create a direct channel from venous to lymphatic pathways – often considered the outflow rout of the glymphatic system – to enhance metabolic waste clearance.^9^

The glymphatic system operates through a three-phase process: (1) CSF influx into the periarterial space, (2) CSF-ISF convection driven by arterial pulsations and aquaporin-4 (AQP4) water channels, and (3) ISF efflux through perivenous spaces toward lymphatic drainage^1,2^. Within this framework, the role of ventricular CSF is critical yet often underemphasized. Ventricular CSF, produced predominantly by the choroid plexus, provides the initial fluid volume and driving pressure necessary for subsequent glymphatic circulation. Its flow dynamics— regulated by anatomical structures and physiological states such as sleep and wakefulness— directly influence the distribution of CSF into periarterial spaces, thus affecting the efficiency of the overall glymphatic clearance pathway. Despite its significance, many aspects of glymphatic physiology remain poorly understood, with ongoing debates regarding its driving forces, efficiency in humans, and its alterations in aging and disease^10,11^. Age-related changes, including vascular stiffening, AQP4 mislocalization, and alterations in CSF production, have been linked to glymphatic dysfunction and may underlie the increased risk of neurodegenerative diseases in older individuals^2,12,13^.

Accurate and clinically feasible measures of glymphatic function, therefore, are essential for assessing metabolic waste removal efficiency and monitoring therapeutic interventions designed to enhance brain fluid clearance, such as LVA.

This further underscores the need for a clinically feasible technique to quantify glymphatic function and assess the efficacy of interventions designed to restore or enhance fluid clearance. Current human imaging approaches primarily rely on magnetic resonance imaging (MRI) techniques, including dynamic contrast-enhanced MRI (DCE-MRI) with intrathecal tracers, phase-contrast MRI (PC-MRI), and diffusion-weighted imaging (DWI), each providing valuable insights into glymphatic flow^8^. However, these MRI methods have notable limitations: DCE-MRI, while valuable for visualizing solute transport, often requires invasive intrathecal administration of gadolinium-based agents, posing risks of discomfort, increased intracranial pressure, and gadolinium deposition in brain tissues^5^. PC-MRI, capable of assessing bulk CSF flow, struggles to detect slower glymphatic dynamics due to limited sensitivity at lower flow velocities^6^. DWI, on the other hand, enables the estimation of molecular diffusion and fluid dynamics^14^, yet its ability to directly assess glymphatic transport remains limited by low spatial resolution and susceptibility to motion artifacts^15^. These technical and practical challenges underscore the critical need for alternative, noninvasive imaging biomarkers. Positron emission tomography (PET), particularly with small-molecule tracers such as 18F-THK and 11C-PiB, has recently been explored for quantifying ventricular CSF clearance as a surrogate of glymphatic function^13,16,17^. Among PET tracers, 18F-fluorodeoxyglucose (18F-FDG) - an analog of glucose - presents a unique opportunity for glymphatic imaging, as preclinical MRI studies using glucose-based tracers have highlighted the relationship between CSF clearance and AD pathology of neurodegenerative diseases such as AD and Huntington’s Disease^18,19^. Unlike receptor-binding PET tracers, 18F-FDG uptake is not confounded by ligand-receptor interactions, allowing for a more direct assessment of CSF dynamics. Furthermore, 18F-FDG is widely available and routinely used in clinical imaging, making it a promising candidate for translational research.

In this study, we propose the use of dynamic 18F-FDG PET to quantify ventricular CSF clearance as a surrogate marker of glymphatic function, leveraging insights from preclinical MRI studies. Our objectives are threefold: (1) to characterize the relationship between ventricular CSF clearance and aging to demonstrate the surrogate markers potential in assessing glymphatic function, (2) to evaluate the reliability of dynamic PET measures using test-retest assessments, and (3) to assess the feasibility of shorter acquisition windows, enhancing clinical applicability. This work aims to establish 18F-FDG PET as a clinically viable tool for indirectly assessing glymphatic function, providing a foundation for future studies on neurodegenerative disease biomarkers and therapeutic interventions.

## Materials and Methods

### Study Population

This study is a secondary analysis of data originally acquired in a randomized clinical trial investigating cerebral glucose metabolism in major depressive disorder^20^. A total of 85 participants (56 females, 29 males; age range 18.1-64.5 years; mean ± SD: 30.3 ± 13.9) underwent baseline 18F-FDG PET imaging. All participants provided written informed consent in accordance with institutional review board guidelines and were compensated for their participation.

Of the 85 baseline scans, five were excluded due to irrecoverable reconstruction issues (e.g. missing attenuation maps or corrupted listmode/normalization files), and additional five were removed due to severe image corruption from excessive motion or instrument error. Three scans were also excluded due to abnormal blood glucose levels (outside 70-150 mg/dL or with fluctuations > 50% between pre- and post-scan). This yielded 72 participants for baseline analysis (Figure 1).

**Figure 1.**
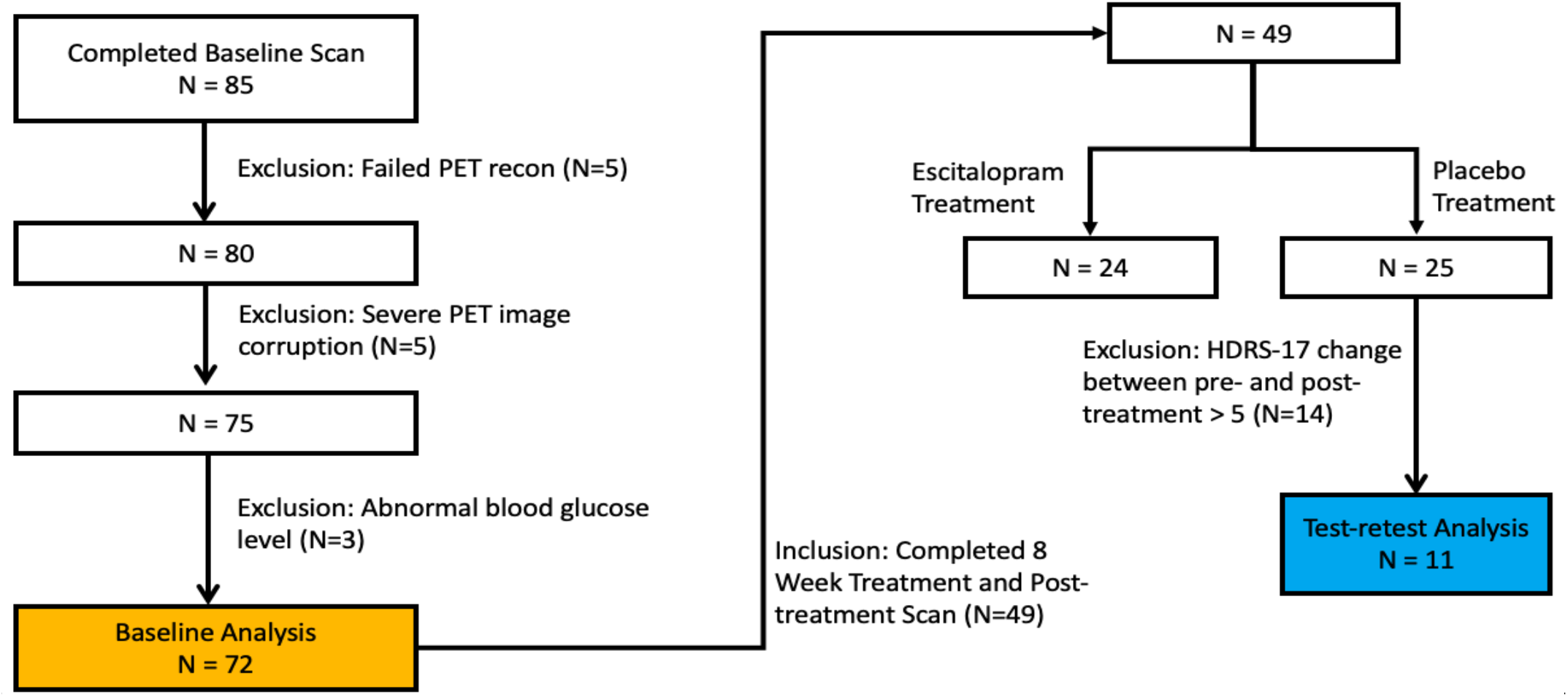
CONSORT-style flowchart detailing participant inclusion and exclusion criteria. A total of 85 individuals completed baseline FDG PET imaging; after quality control and glucose-level screening, 72 were included in baseline analyses. Of these, 49 completed post-treatment scans following an 8-week intervention. For test–retest analysis, only participants from the placebo arm with HDRS-17 change ≤5 were retained (N=11).

Of these, 49 participants completed an 8-week treatment protocol followed by a second/post-treatment PET scan. As part of the trial, participants were randomized to receive escitalopram (N=24) or placebo (N=25). Since there is evidence that escitalopram affects glymphatic function^21^, only participants from the placebo group were considered for test-retest evaluation. Among them, 14 were excluded due to Hamilton Depression Rating Scale (HDRS-17) changes larger than 5 from pre- to post-treatment, resulting in 11 participants for the test-retest analysis (Figure 1).

### PET/MR Data Acquisition

Each participant received an intravenous injection of 148–185 MBq (4–5 mCi) of 18F-FDG and was placed in a Siemens Biograph mMR PET/MR scanner for a continuous 60-minute scan. PET data were acquired in list-mode format throughout the 60-minute session, then binned into thirty 2-minute frames to capture tracer uptake kinetics. Reconstructions were performed using Siemens E7 Tools with a maximum-likelihood expectation maximization (MLEM) algorithm for 75 iterations. We used 75 iterations of the MLEM algorithm based on the contrast-to-noise ratio (CNR) between gray and white matter. CNR plateaued after around 75 iterations, indicating sufficient image quality without further improvement ^22^. Concurrently, high-resolution anatomical MR images were acquired using a T1-weighted MPRAGE (Magnetization Prepared Rapid Gradient Echo) sequence with typical parameters: repetition time = 2300 ms, echo time = 2.98 ms, inversion time = 900 ms, flip angle = 9°, 1 mm isotropic resolution. MR-based attenuation correction was applied using the “Boston method,”^23^ which uses tissue segmentation and atlas registration to model soft tissue and bone structures. Participants were instructed to remain still and awake throughout the scan, and no sedation was used. The resulting co-registered PET and T1-weighted MR data provided the basis for subsequent analyses of glymphatic clearance dynamics.

### Images processing

The 30 dynamic PET frames were first averaged to create a composite mean PET image, which served as a reference for motion correction. Each original frame was rigidly realigned (six degrees of freedom) to this composite mean. Then the same averaging and realignment steps were repeated once more to minimize residual head motion. After motion correction, the aligned PET frames were co-registered to the corresponding T1-weighted MR image using Freesurfer 7.4.1. Subsequently, for each subject, the radioactivity concentration of all 30 frames was normalized by body weight and injected dose to generate standardized uptake value (SUV) images. A subject-specific region of interest (ROI) encompassing the lateral ventricles was extracted using FreeSurfer, which was then eroded using a single-voxel morphological erosion to reduce partial-volume effects from surrounding tissues. Time-activity curves (TACs) were finally extracted from the eroded ventricular ROI across all dynamic frames and used for subsequent kinetic modeling and analyses.

### Modeling of the Time-Activity Curves

To model the dynamic behavior of 18F-FDG in the lateral ventricles, we employed the γ-variate model^24^, which provides a physiologically plausible representation of bolus kinetics. As a glucose analog, FDG exhibits distribution and clearance characteristics similar to native glucose following intravenous injection. Prior work using dynamic glucose-enhanced MRI has demonstrated that the γ-variate model accurately captures the temporal profile of glucose concentrations in both brain parenchyma and CSF, including the initial rise and subsequent clearance phase. Using this model, the TAC of the lateral ventricle can be modeled as:

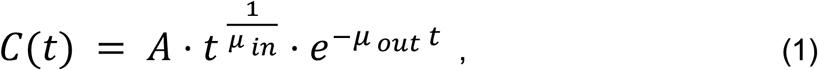

where C(t) is the standardized uptake value (SUV) at time t, A represents the overall amplitude, *μ*_*in*_ represents the 18F-FDG influx rate into the ventricles, and *μ*_*in*_ indicates the CSF clearance (efflux) rate, respectively. In general, *μ*_*in*_ quantifies how fast the TAC curve rises and *μ*_*out*_ quantifies how fast the TAC curve drops. Note that the two variables do not act independently but jointly determine the shape of the curve.

### Test-retest analysis

To assess the stability of ventricular CSF clearance measures, we performed a test-retest analysis in N=11 participants (Figure 1) who received placebo and completed both baseline and 8-week follow-up PET scans. The 8-week interval was deemed insufficient for aging-related effects to impact glymphatic dynamics. Only participants with no larger than 5-point change on the HDRS-17 from pre- to post-treatment were included, consistent with the minimal clinically important difference^25^, to minimize confounding effects from clinically meaningful mood changes. The exclusion of large HDRS-17 change is due to the association between depression level and CSF clearance, which will be justified in the Results section. The same image preprocessing and modeling pipeline was applied at both time points. Fitted *μ*_*out*_ values were compared to evaluate test-retest reliability and repeatability.

### Feasibility Analysis of Reduced Scan Duration

To evaluate the clinical feasibility of shorter imaging protocols, we investigated whether scanning durations of less than one hour could still capture reliable information about CSF clearance. Beginning at 10 minutes post-injection, we varied the analysis window by adjusting the end time, with each interval spanning a minimum of 10 minutes. For each start–end time window [*t*_*start*_, *t*_*end*_ ], a TAC was extracted from the eroded lateral ventricle ROI. A linear model was fitted to the SUV values within the interval to estimate a clearance slope. This slope was then normalized by the mean SUV of the same interval to enable inter-subject comparison. Thus, the normalized clearance slope between start time *t*_*start*_ and end time *t*_*end*_ is:

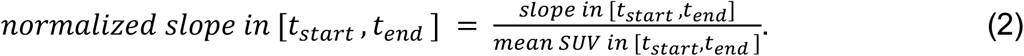

Correlation analysis was performed between the normalized slopes and both the efflux rate (*μ*_*out*_) and participants’ age across all 72 baseline scans. By tracking how these correlations varied across different start-end time windows, we identified shortened scan durations that preserved the sensitivity to individual differences in CSF clearance.

### Statistical Analysis

The primary outcome measures, *μ*_*in*_*(influx rate) and* *μ*_*out*_*(efflux rate)*, were first examined for normality using visual inspection and the Shapiro-Wilk test. As both variables conformed to normality assumptions, their associations with age were assessed using Pearson correlation.

To account for potential confounders, separate multiple linear regression models were constructed for *μ*_*in*_ and *μ*_*out*_, incorporating age, lateral ventricle size, baseline HDRS-17 score, and sex, as predictors.

Test–retest reliability was assessed in a subset of N=11 participants. The intraclass correlation coefficient (ICC) was calculated to quantify the reproducibility of *μ*_*out*_ across the two scan sessions.

For the feasibility analysis of reduced scan duration, Pearson correlation test was used to assess the relationship between normalized clearance slopes (derived from truncated TAC intervals) and both *μ*_*out*_ and age. Correlation strength was tracked across varying start–end time windows to evaluate the reliability of shortened imaging protocols.

All statistical tests were two-tailed, with significance defined as p < 0.05, unless otherwise specified. Analyses were performed using SPSS 29.0.2, and figures were generated with Python 3.11 using matplotlib 3.7.2.

## Results

### Baseline Analysis: Association Between Ventricular CSF Clearance and Age

To investigate age-related differences in ventricular CSF clearance, we analyzed 72 baseline (pre-treatment) FDG PET scans, minimizing potential confounds from treatment or placebo effects. Demographic characteristics by sex are summarized in Table 1. Although females were slightly older on average than males, no significant sex differences were observed in baseline depression severity (HDRS-17) or lateral ventricle size.

**Table 1.** Baseline Participant Demographics.

| Sex | N | Age | HDRS-17 | Lateral Ventricle Size (cc) |
| --- | --- | --- | --- | --- |
| M | 25 | 29.6 (11.8) | 18.2 (4.8) | 14.86 (6.24) |
| F | 47 | 30.9 (15.3) | 17.3 (4.4) | 12.20 (5.63) |
|  |  | P = 0.034* | P = 0.536 | P = 0.286 |

Values are expressed as mean (SD). Males and females are compared using independent two-sample t-test (\* $p < 0.05$ ). HDRS-17: Hamilton Depression Rating Scale – 17.

Figure 2 displays representative dynamic PET images and fitted time-activity curves (TACs) with the γ-variate model for a young participant in early adulthood and an older participant in their 60s. In both cases, 18F-FDG progressively accumulates in the lateral ventricles, but notable differences in curve shape are evident. The younger participant’s TAC plateaus and declines after ∼30 minutes, yielding a positive *μ*_*out*_ value consistent with intact CSF clearance. In contrast, the older participant’s TAC continues to rise throughout the scan, producing a negative *μ*_*out*_ value indicative of impaired clearance.

**Figure 2.**
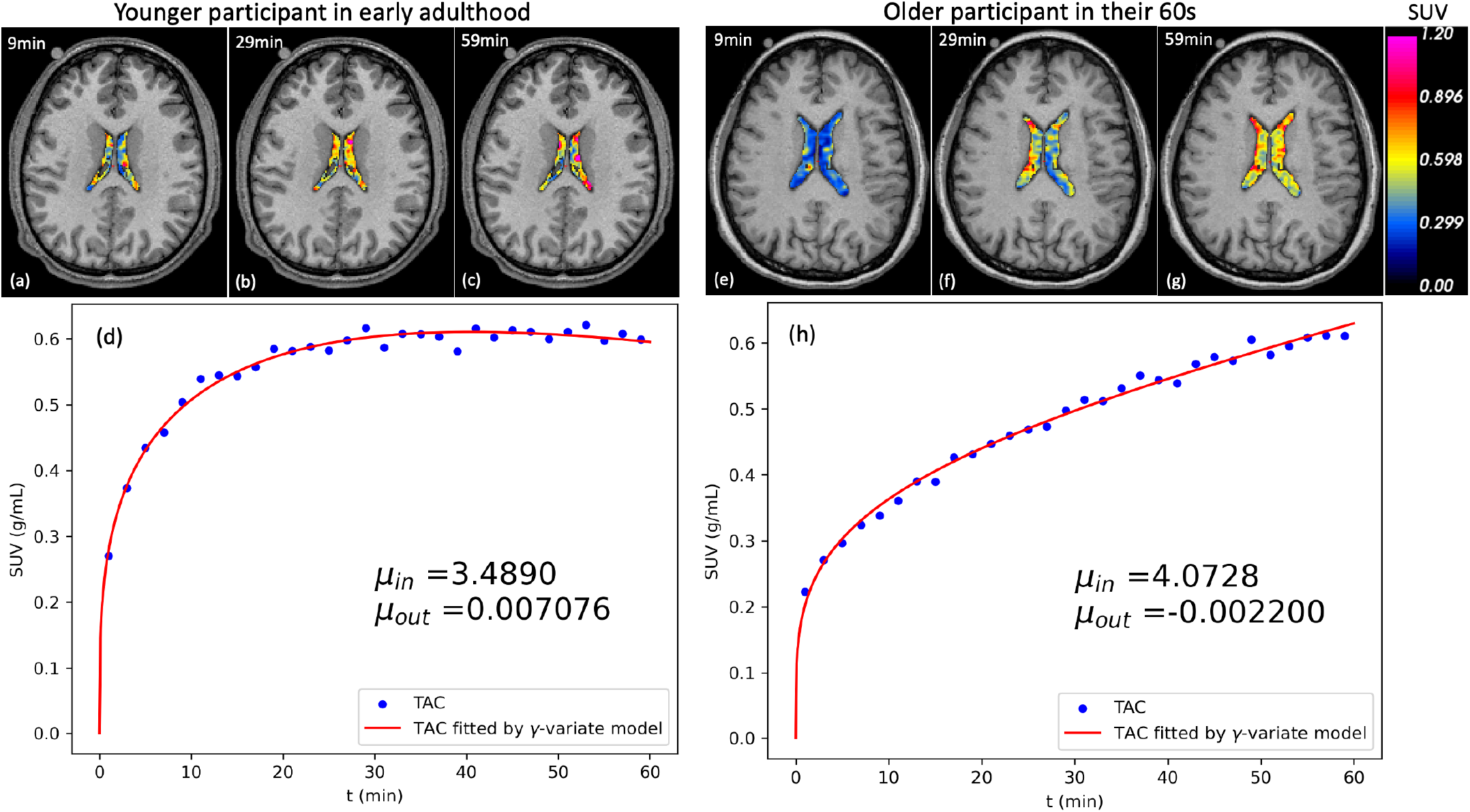
Representative dynamic PET/MR images (a, c) and corresponding lateral ventricle time-activity curves (TACs; b, d) (b, d) for one younger participant and one older participant. The influx (*μ*_*in*_) and clearance (*μ*_*out*_) rates were estimated using γ-variate model. The younger participant’s curve plateaus and declines after ∼30 minutes, indicating effective CSF clearance (positive *μ*_*out*_), whereas the older participant’s curve continues to rise throughout the scan, yielding a negative *μ*_*out*_ consistent with impaired clearance.

To evaluate these patterns across the cohort, lateral ventricle TACs were normalized by their 2-norm (Figure 3a) and fitted by the γ-variate model (Figure 3b). A clear age-related gradient was observed: older participants (depicted in red/yellow) exhibited flatter, continuously rising curves, while younger individuals (blue/green) showed earlier plateaus suggestive of more efficient CSF clearance.

**Figure 3.**
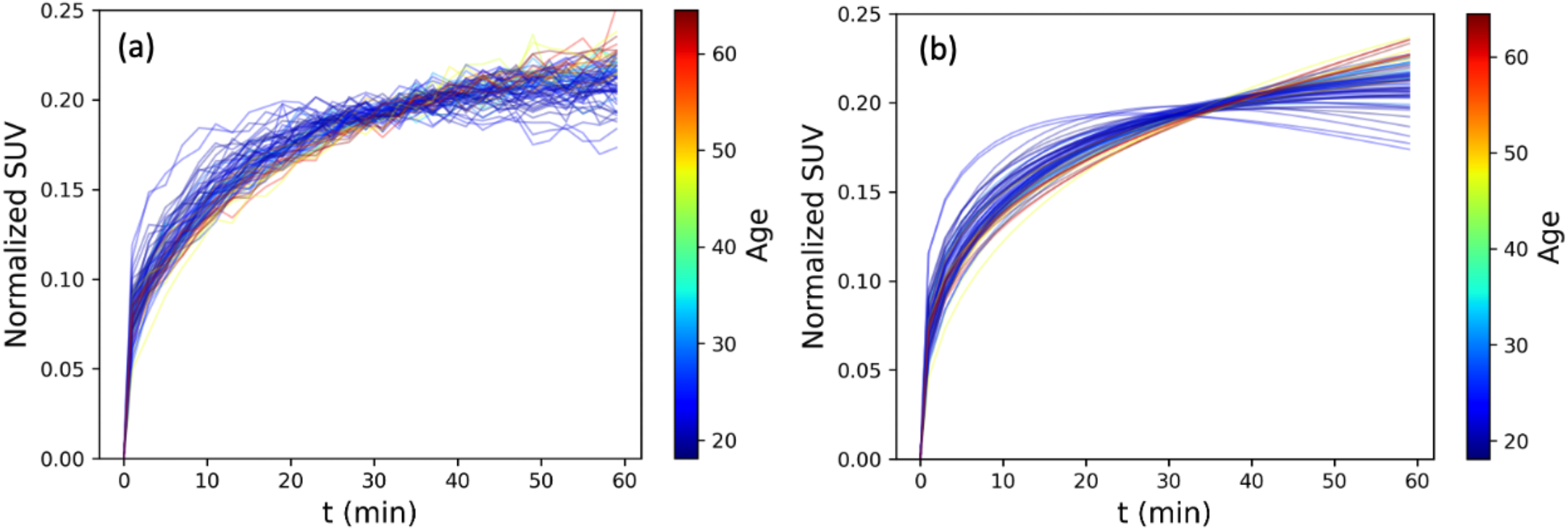
Normalized lateral ventricle TACs (a) and corresponding γ-variate model fits (b) for all 72 participants. Curves are color-coded by age.

We then quantified the relationship between fitted parameters and age. As shown in Figure 4, *μ*_*in*_ was not significantly associated with age (r = 0.203, p = 0.087; Fig. 3a), whereas *μ*_*out*_ exhibited a strong negative correlation (r = –0.680, p < 0.001; Fig. 3b), indicating that CSF clearance declines with age while influx remains stable.

**Figure 4.**
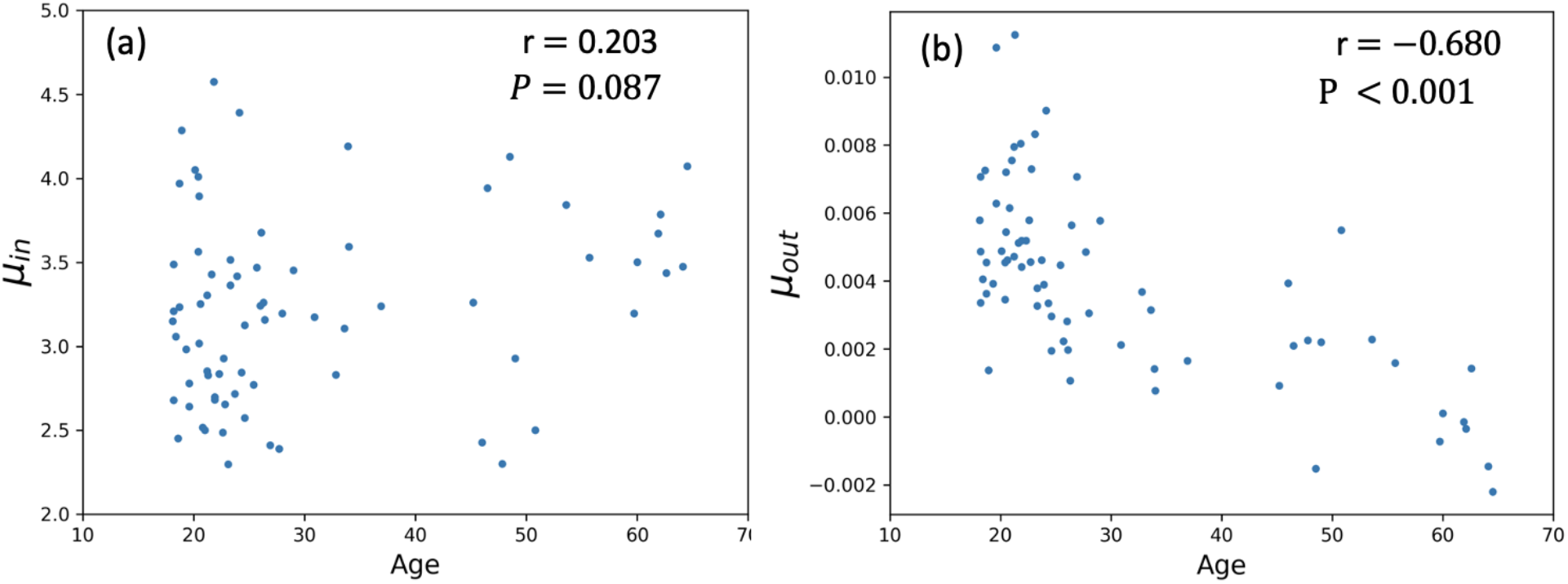
Scatter plot of fitted parameters (a) *μ*_*in*_ and (b) *μ*_*out*_ of the γ-variate model to the subjects’ TACs against age, along with Pearson correlation test results.

To further assess whether lateral ventricle size, baseline HDRS-17 score or sex influenced CSF clearance, we performed a multiple linear regression analysis. Table 2 shows the analysis results. For *μ*_*out*_, age emerged as the strongest predictor (standardized beta = -0.584, p < 0.001), and baseline HDRS-17 was also significant (standardized beta = 0.200, p = 0.021), whereas ventricle size approached borderline significance (standardized beta = -0.191, p = 0.052) and sex showed no association (p = 0.956). In contrast, analysis of *μ*_*in*_ revealed significant effects for both age (standardized beta = 0.361, p = 0.006) and lateral ventricle size (standardized beta = -0.362, p = 0.007), but no significant associations with baseline HDRS-17 (p = 0.289) or sex (p = 0.160). These findings confirmed age as a robust predictor of both influx (*μ*_*in*_) and efflux (*μ*_*out*_) parameters. Depression severity (as captured by HDRS-17) is selectively linked to efflux that higher *μ*_*out*_ was associated with more severe depression. Lateral ventricle size may also modulate these CSF dynamics.

**Table 2.**
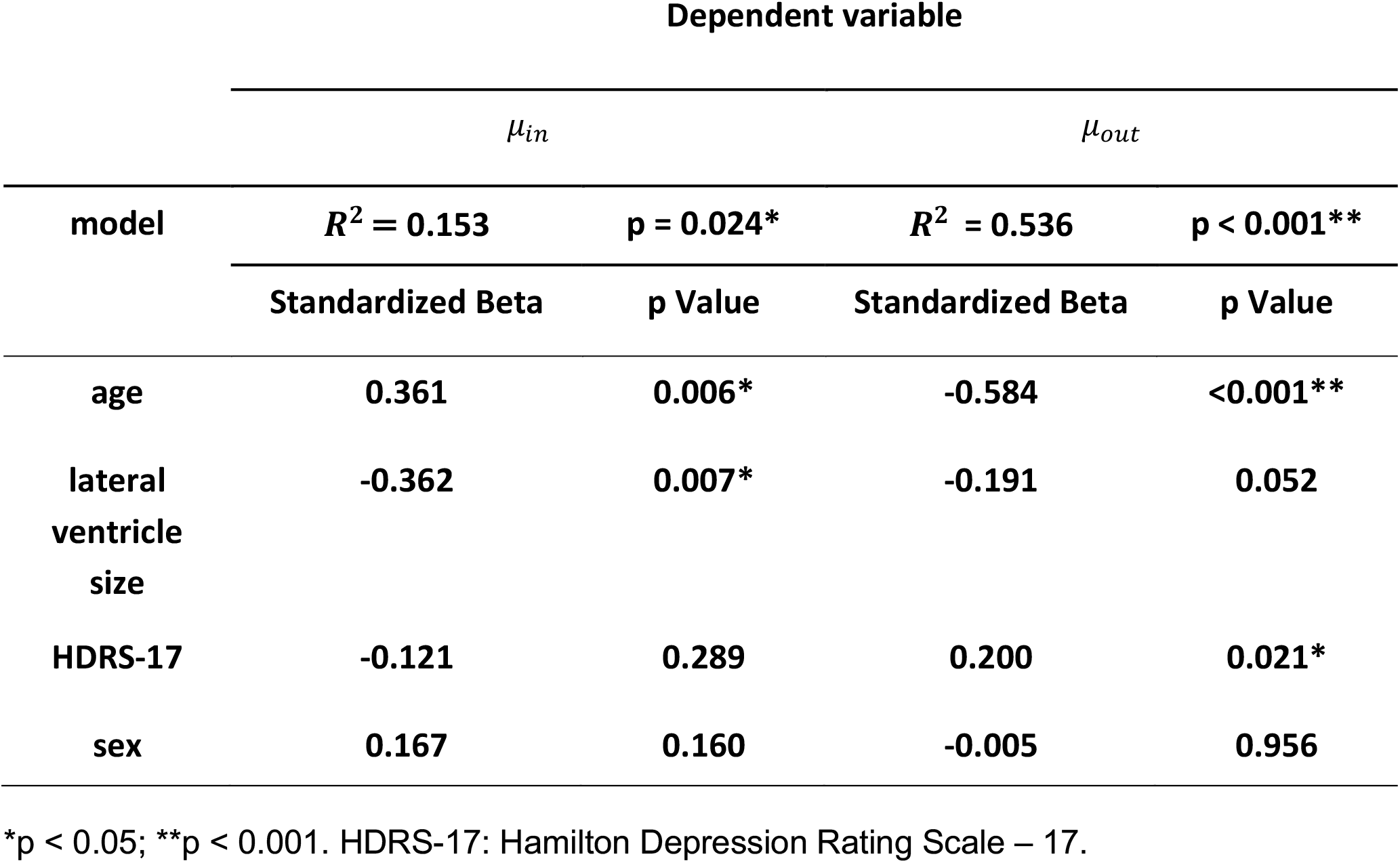
Multiple linear regression results for influx rate *μ*_*in*_ and clearance rate *μ*_*out*_. Covariates included age, lateral ventricle size, HDRS-17 score, and sex.

To further explore the influence of depression severity, we regressed out age, lateral ventricle size, and sex from *μ*_*out*_ and correlated with the residuals with HDRS-17 (Figure 5). A significant positive association remains (r = 0.276, p = 0.019), indicating the independent influence of depression severity on CSF clearance.

**Figure 5:**
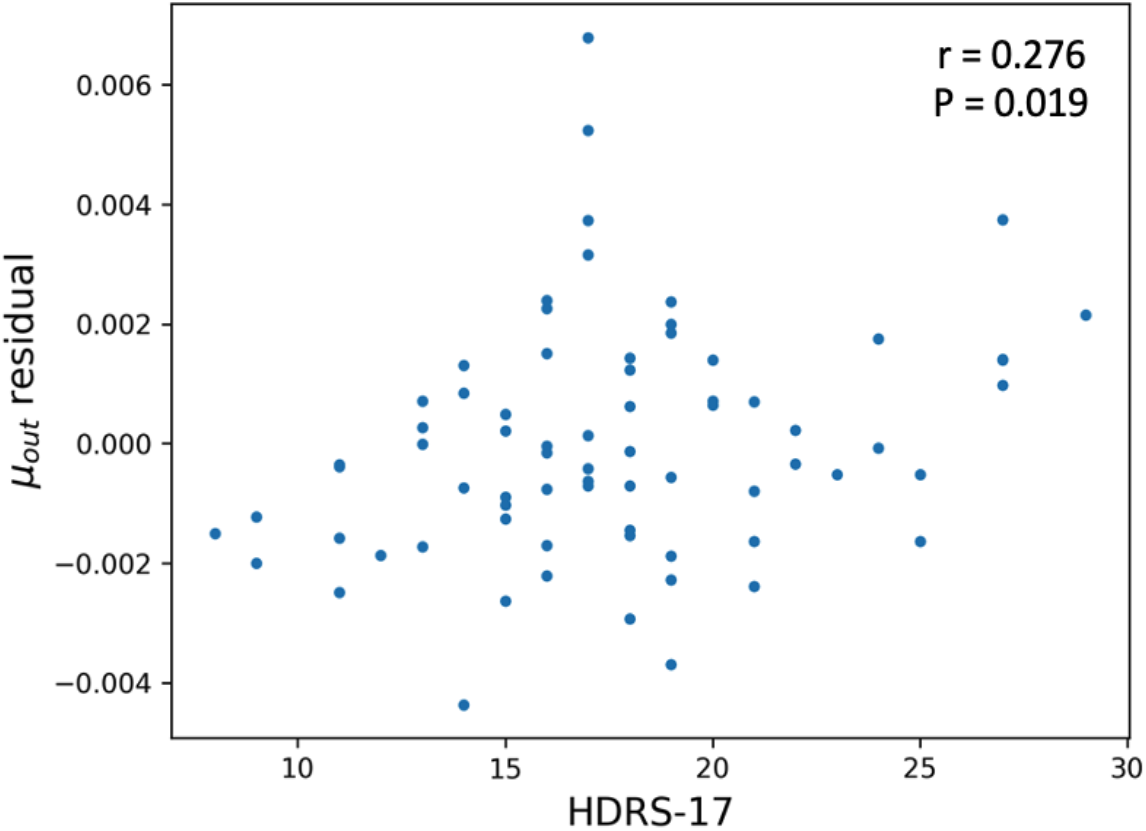
Relationship between HDRS-17 scores and *μ*_*out*_ residuals after adjusting for age, lateral ventricle size, and sex through multiple linear regression (Table 2). Pearson correlation test result is displayed.

### Test-retest analysis

We next assessed the reliability of *μ*_*out*_ in a subset of 11 participants who received placebo and exhibited minimal change in HDRS-17 scores (<5 points) between baseline and post-treatment scans. Figure 6 shows the comparison of *μ*_*out*_ values across timepoints, with an intraclass correlation coefficient (ICC) of 0.702. This indicates moderate to good test–retest reliability of ventricular CSF clearance measured by *μ*_*out*_.

**Figure 6.**
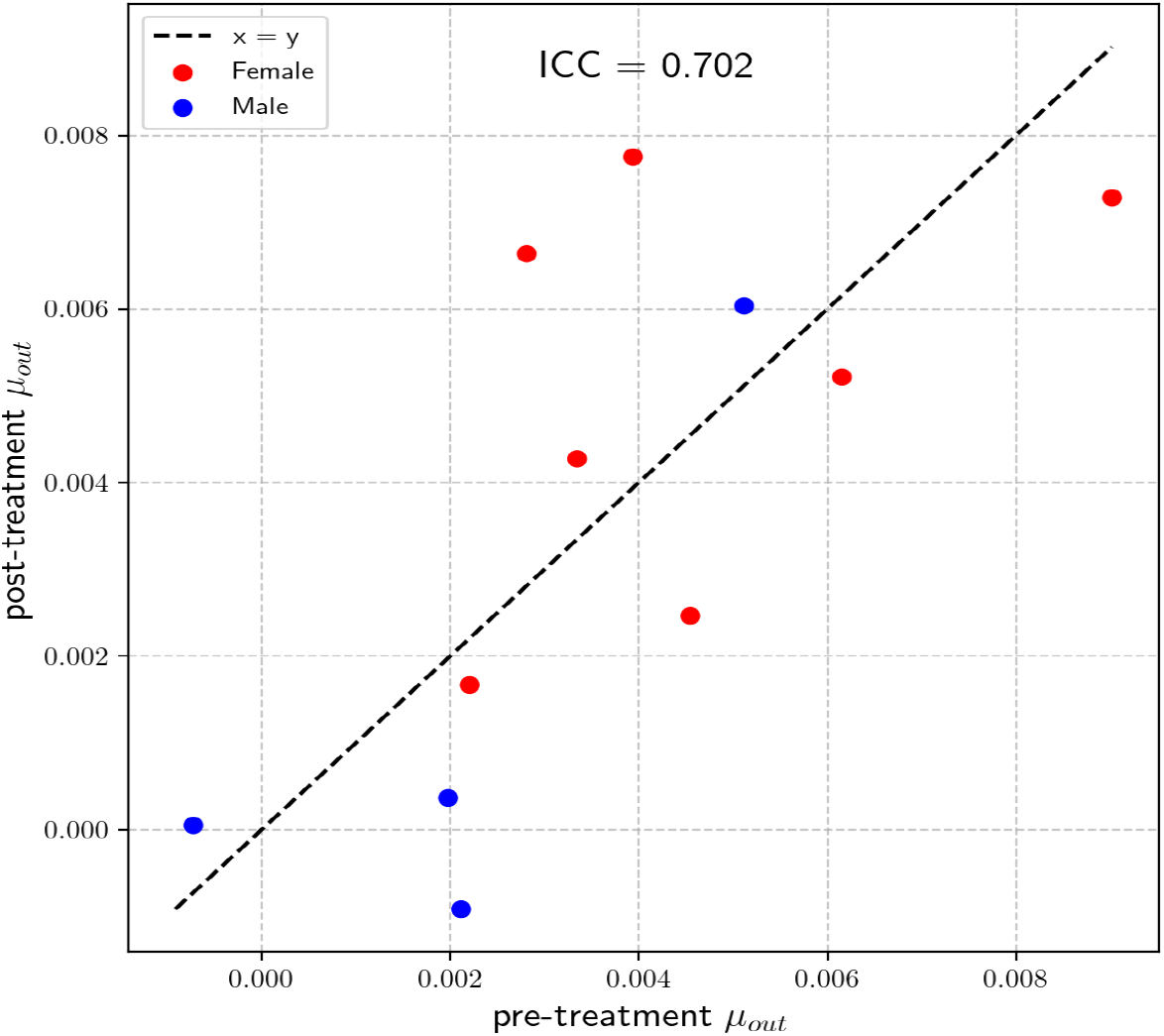
Test–retest reliability of *μ*_*out*_ across two scans spaced 8 weeks apart in N=11 placebo-treated participants. Dashed line represents identity. ICC (intraclass correlation coefficient) = 0.702.

### Feasibility Analysis of Reduced Scan Duration

To evaluate the potential for shorter PET imaging protocols, we examined whether clearance metrics could be preserved using truncated scan windows. For each pair of start and end times (minimum 10 minutes apart) beginning at 10 minutes post-injection, we fit a linear model to the ventricular TAC and normalized the slope by the mean SUV over the interval.

Figures 7a and 7b display the Pearson correlation between these normalized slopes and both *μ*_*out*_ and age across varying scan windows. Correlation strength increased with longer scan durations, peaking when the end time approached 60 minutes.

**Figure 7.**
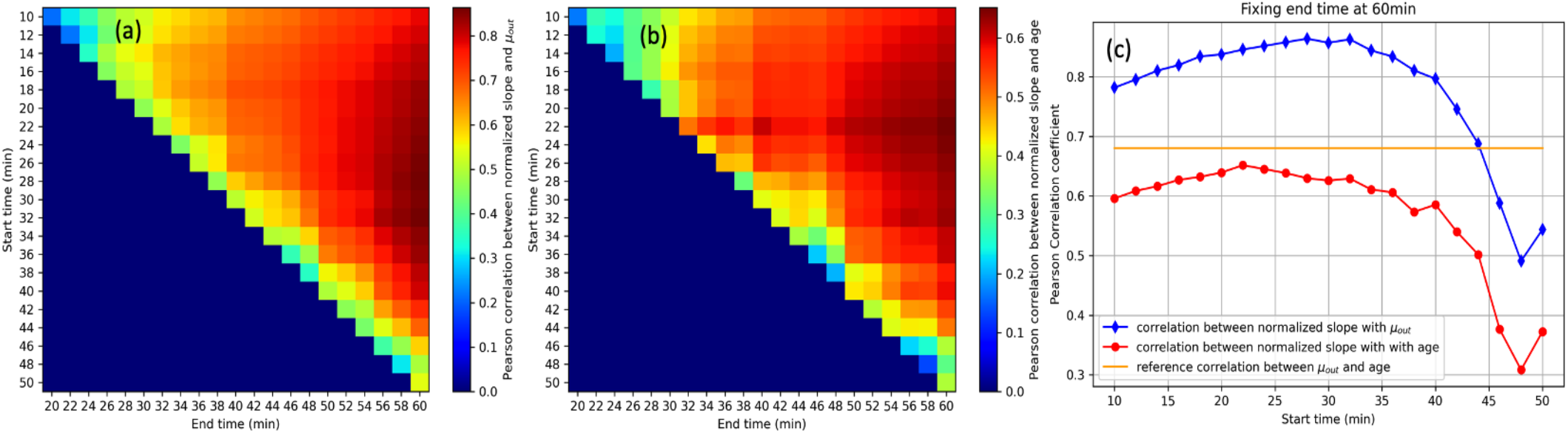
Evaluation of reduced scan duration. Correlation map of normalized slope of a shortened TAC versus (a) *μ*_*out*_ and (b) age, across various scan (y-axia) start time and (x-axis) end time. (c) Correlation curves between the normalized slope and *μ*_*out*_ (and age), with fixed end time at 60 minutes.

Fixing the end time at 60 minutes and varying the start time (Figure 7c) showed that correlations with *μ*_*out*_ increased up to ∼30 minutes and then declined, while correlations with age remained stable until ∼30 minutes and then dropped sharply. This suggests that a 30–60 minute scan window preserves maximal association with both *μ*_*out*_ and age.

These results demonstrate that a reduced protocol—from 30 to 60 minutes post-injection— could capture essential CSF clearance dynamics with minimal loss of sensitivity, offering a feasible path toward abbreviated clinical imaging strategies.

## Discussion

There findings provide direct in vivo evidence that ventricular CSF clearance decreases with advancing age, consistent with earlier preclinical MRI studies demonstrating that aging slows CSF turnover and impairs metabolic waste elimination^26^. As illustrated in Figures 2 and 3, older participants exhibited reduced 18F-FDG clearance in the lateral ventricles, suggesting that the glymphatic function—of which CSF flow is a major driving component—may be compromised in later life. This observation aligns with previous reports of reduced ISF flow after age 50^27^, potentially driven by factors such as reduced arterial pulsatility and AQP4 mislocalization^28^. Multiple linear regression analysis (Table 2) indicated that, although older individuals tend to have structurally enlarged ventricles^29^, age-related clearance decline could not be explained by this morphological feature. Nor does sex appear to play a significant role. These results support the notion that age-related deficits in glymphatic clearance reflect a physiological process beyond simple structural change.

Compared to existing MRI-based glymphatic imaging techniques, our PET-derived measure of ventricular CSF clearance offers several notable advantages. As discussed in the Introduction, PC-MRI is well-suited for quantifying high-velocity bulk flow but struggles to reliably detect slow-moving CSF within small anatomical regions like the ventricles. Extended MRI acquisition times required to enhance signal-to-noise ratio also pose logistical difficulties, especially for older adults and cognitively impaired individuals. DCE-MRI methods, which utilize intrathecal administration of gadolinium-based contrast agents, are invasive, time-consuming, and may alter physiological conditions due to higher molecular-weight tracers. By contrast, PET imaging with the widely available 18F-FDG tracer circumvents these limitations. It requires only standard intravenous injection and accommodates a practical scanning duration suitable for clinical populations. Crucially, the relatively long half-life and minimal binding complexity of 18F-FDG allow for accurate detection of subtle, prolonged clearance patterns characteristic of impaired glymphatic function in aging populations. Together, these attributes position PET imaging as a valuable alternative or complementary approach to existing MRI techniques, effectively addressing critical gaps related to invasiveness, patient comfort, logistical feasibility, and sensitivity to subtle glymphatic alterations.

Test-retest analysis (Figure 6) showed that *μ*_*out*_ exhibited moderate-to-good reliability (ICC = 0.702) over an 8-week interval in placebo-treated participants with stable clinical status. Although the sample size was limited (N=11), this degree of reproducibility supports *μ*_*out*_ as a reliable CSF clearance marker for longitudinal monitoring, potentially enabling detection of changes over time in disease progression or response to therapeutic intervention.

One observation from our study is that many participants, particularly older ones, displayed a continuously rising TAC at the 60-minute mark, suggesting that a one-hour scan captures only the early phase of CSF clearance. This observation implies that clearance dynamics may be prolonged in older adults, and longer scans could be necessary to fully characterize tracer outflow. While the 60-minute window provides useful information about CSF clearance kinetics, extended observation may yield more robust estimates of *μ*_*out*_—particularly in detecting subtle or protracted changes among older adults. If future studies employ longer imaging sessions, it may be possible to differentiate more definitively between normal and compromised ventricular clearance rates in humans, thereby enhancing the clinical utility of 18F-FDG PET for glymphatic assessment.

Interestingly, *μ*_*out*_ remained positively associated with HDRS-17 scores after controlling for age, ventricle size, and sex (Figure 5), indicating that individuals with more severe depressive symptoms may exhibit higher CSF clearance rates. While the underlying mechanisms remain speculative, one possibility is that elevated stress-related hormones or altered neurovascular regulation in depression could transiently augment CSF outflow. Alternatively, this correlation may be caused by a compensatory mechanism in which the brain attempts to clear potentially harmful metabolic byproducts more rapidly under heightened psychiatric distress. However, the direction and causality of this relationship remain unclear, warranting future studies that incorporate additional physiological and neuroendocrine measures to clarify underlying mechanisms.

Our feasibility analysis demonstrated that a truncated 30-minute window starting at 30-minutes post injection captured much of the information contained in the full 60-minute scan. Normalized slope estimates from this window remained strongly correlated with both *μ*_*out*_ and age (Figure 7). These findings support the potential use of abbreviated scanning protocols, which holds promise for clinical applications and may be particularly beneficial in older or clinically populations who may struggle with long acquisition times. Future studies that validate the reproducibility and diagnostic value of such abbreviated protocols in larger, more diverse populations and longer acquisition time will help determine whether a reduced scan duration could become a standard practice in assessing CSF clearance.

Specifically, the strength of 18F-FDG PET in assessing CSF clearance lies in its relatively low binding complexity compared to other PET tracers that have been employed for a similar purpose. Tau radioligands (e.g., 18F-THK5117^16^, 18F-THK5351^13^), amyloid-beta agents (e.g., 11C-PiB^17^), and inflammation-targeting tracers (e.g., 18F-FEPPA^30^) all interact with specific binding sites, introducing complexities due to variations in receptor expression, affinity, and tissue uptake kinetics. As a result, these tracers often exhibit a rapid drop in blood activity during the early phase, and their late-phase interpretation is confounded by multipass blood supply, binding competition, and transporter limitations. Furthermore, for extended studies (∼120 minutes), the residual tracer signal in the ventricles may become insufficient to clearly distinguish inter-individual differences in clearance rates. By contrast, 18F-FDG combines the advantages of an 18F label with a 110-minute half-life, a widely available clinical protocol, and the absence of a receptor-binding step, which simplifies kinetic modeling of ventricle outflow over an extended timeframe. Moreover, 18F-FDG is a glucose analog, making it directly comparable to preclinical studies that use glucose-based tracers to evaluate CSF and glymphatic dynamics. While non-binding, water-like tracers such as 11C-butanol^31^ or 15O-H2O^32^ circumvent receptor interactions, their very short half-lives (20 and 2 minutes, respectively) make prolonged imaging impractical. Taken together, these attributes make 18F-FDG a practical and potentially more robust tracer for quantifying CSF dynamics and evaluating glymphatic function in both research and clinical settings.

This work’s generalizability is limited by the sample, drawn exclusively from individuals with major depressive disorder. Although convenient for secondary analysis of age-related changes, depression itself — with its associated physiological and metabolic alterations — may influence CSF clearance and glymphatic function. In fact, we observed a positive correlation between depression severity and *μ*_*out*_ in this study (Figure 5). Prospective studies including healthy controls and individuals with diverse neuropsychiatric profiles are needed.

In addition, the γ-variate model—while physiologically grounded—assumes a single bolus-shaped clearance trajectory, which may oversimplify the multi-compartmental nature of CSF movement and the multiphasic behavior of glymphatic dynamics, especially over extended time periods or in pathological conditions. It may also fail to account for partial reabsorption over longer intervals, as well as the kinetic nuances introduced when FDG is phosphorylated and becomes trapped intracellularly. Future modeling efforts may benefit from compartmental or multiphasic frameworks to better capture these complexities.

## Conclusion

This study used dynamic 18F-FDG PET imaging to quantify ventricular CSF clearance, which is shown to decline with age, implying impaired glymphatic function in later life. Using a γ-variate model, we demonstrated that the CSF clearance quantification is reliable and reproducible. Our results suggest that even abbreviated imaging protocols – focusing on the 30-60 minute post-injection window – can potentially preserve key clearance metrics, facilitating broader clinical application. Compared to existing MRI-based approaches, 18F-FDG PET offers practical, noninvasive advantages for assessing ventricular CSF circulation. Future studies incorporating longer scan sessions and healthy control populations are needed to further validate this approach. Additionally, exploring more advanced models may help better capture the complex kinetics of ventricular tracer dynamics. Together, these findings support the use of dynamic 18F-FDG PET as a promising tool for investigating glymphatic function in aging and neuropsychiatric conditions, with potential implications for early detection, monitoring, and therapeutic targeting of brain clearance deficits.

## Disclosure

Data used in this study were originally acquired with the support of NIH/NIMH grant R01MH104512 (CD). The authors declare no competing interests. No financial relationships, activities, or affiliations exist that could be perceived as having influenced the submitted work beyond the stated funding support.

## Data Availability Statement

All data produced in the present study are available upon reasonable request to the authors

